# Effects of counteracting measures on muscle contraction determinants in CKD: what muscle biopsy studies tell us?

**DOI:** 10.1101/2022.12.30.22284067

**Authors:** Georgia I. Mitrou

## Abstract

Chronic kidney disease (CKD) is characterized by muscle atrophy, fatigue intolerance and other indicators of muscle dysfunction, collectively termed uremic myopathy, with devastating consequences in overall health status and mortality rates. Although many factors such as metabolic acidosis, substrate availability and neuropathy have been implicated, the mechanisms underlying uremic myopathy have not yet been fully understood. However, there is clear evidence that muscle specific factors such as fiber atrophy, fiber type alterations and mitochondrial abnormalities are presented in muscle biopsies of CKD patients and can negatively affect muscle contraction. Counteracting measures such as exercise and nutritional interventions have been shown to improve muscle performance, health indices and overall quality of life of CKD patients. However, little is known about their effects on factors affecting muscle contraction at the muscle biopsy level and therefore on the mechanisms underlying uremic myopathy. The current systematic review aims to summarize the effects of recent interventional studies on muscle contraction determinants based on muscle biopsies of human patients.

## Introduction

Chronic kidney disease (CKD) is increasingly recognized as a major global health problem affecting more than 10% of global population and is considered as one of the leading causes of death worldwide (1). CKD patients present with skeletal muscle structural and functional abnormalities (2–4) with symptoms of muscle weakness, limited endurance and fatigue intolerance (5), neuropathy (6) and a host of other striated muscle problems, collectively termed *uremic myopathy* (5). Uremic myopathy is a common condition in CKD patients which in turn leads to high morbidity and mortality rates and worsens in patients with end-stage renal disease (7).

Many factors have been implicated in uremic myopathy including metabolic acidosis, substrate availability and neuropathy (6). A host of confounding factors such as years from diagnosis, comorbidities, pharmaceuticals and nutritional status add more complexity in understanding the mechanisms underlying uremic myopathy. However, muscle specific factors such as fiber atrophy (8–10), fiber type alterations (11) and abnormalities in mitochondrial form and function (12,13), have been reported in muscle biopsies of CKD patients and can negatively affect muscle contraction. Recent studies have also reported additional muscle abnormalities at the muscle biopsy level including oxidative/nitrosative stress (14,15), inflammation (16,17), fat accumulation/infiltration (16,17) and fibrosis (18,19) which may also affect muscle contraction. Moreover, a previous study from our group (20) in an animal model of CKD, reported that muscle dysfunction of single muscle fibers, is not fully explained by atrophy (11% smaller fiber areas compared to control fibers). Specifically, a significantly lower maximal isometric force was observed in CKD muscle fibers compared to controls even after correcting the values for the observed atrophy. This indicates muscle specific impairments in uremic myopathy which are independent of other factors such as neuropathy and substrate availability.

Numerous studies have examined the effects of various interventions (pharmaceutical, non-pharmaceutical or combination) to improve muscle performance, health indices and overall quality of life and mortality rates in CKD patients. However, little is known about their effects on muscle contraction determinants on skeletal muscle biopsies of CKD patients. The aim of the current systematic review is to summarize the effects of recent interventional studies on skeletal muscle biopsies of CKD patients aiming to understand any aspect of skeletal muscle quality and quantity which could be affected. This could help us to better understand the mechanisms underlying uremic myopathy and therefore to support the design of appropriate interventions in order to prevent or alleviate its devastating impacts.

## Methods

The current review includes original research articles studying muscle biopsies from CKD patients (including patients undergoing hemodialysis or peritoneal dialysis) before and after any intervention. Pubmed and Scopus were searched between March and April 2022 by two reviewers according to the Cochrane (Higgins and Green, 2011) and PRISMA (Moher et al., 2009) guidelines with the keywords ‘((((chronic kidney disease) OR chronic renal failure) OR end-stage renal disease) OR uremia) AND skeletal muscle AND ((muscle function) OR muscle contraction) AND (((muscle cell) OR muscle fiber) OR muscle fibre)’. In order to limit the results in articles published in the last ten years, written in English and in studies assessing human samples, the filters ‘publication date (10 years)’, ‘language (English)’ and ‘species (humans)’ were applied. The articles were selected according to inclusion and exclusion criteria. We included studies assessing skeletal muscle biopsies from CKD patients in comparison to their baseline values (and control subjects when available). Articles which assessed other than skeletal muscle biopsies, muscle biopsies from other clinical populations or CKD patients after transplantation were excluded from this review. Moreover, case reports, reviews, guidelines and studies without control group were also excluded.

### Overall Results

This review includes original studies assessing muscle contraction determinants based on muscle biopsies of CKD patients (all stages). Therefore, in Pubmed and Scopus the literature search identified 111 and 149 articles respectively. After removing duplicates (50), 210 articles remained for screening. Of those, 4 studies met the inclusion criteria **(Figure 1)** and 206 articles were excluded for the following reasons: 15 articles had assessed other tissues than skeletal muscle biopsies, 19 articles had assessed muscle biopsies from CKD patients but not before and after any intervention, 30 articles had assessed other than CKD patients, 3 articles had assessed CKD patients after transplantation, 104 articles were reviews, 2 articles were case reports and 7 articles were other than research articles (test, comment, editorial, guidelines and abstract). Moreover, although the filter ‘species (humans)’ was applied, the literature search identified 26 studies which had used animal models which were also excluded.

**Figure 1.**
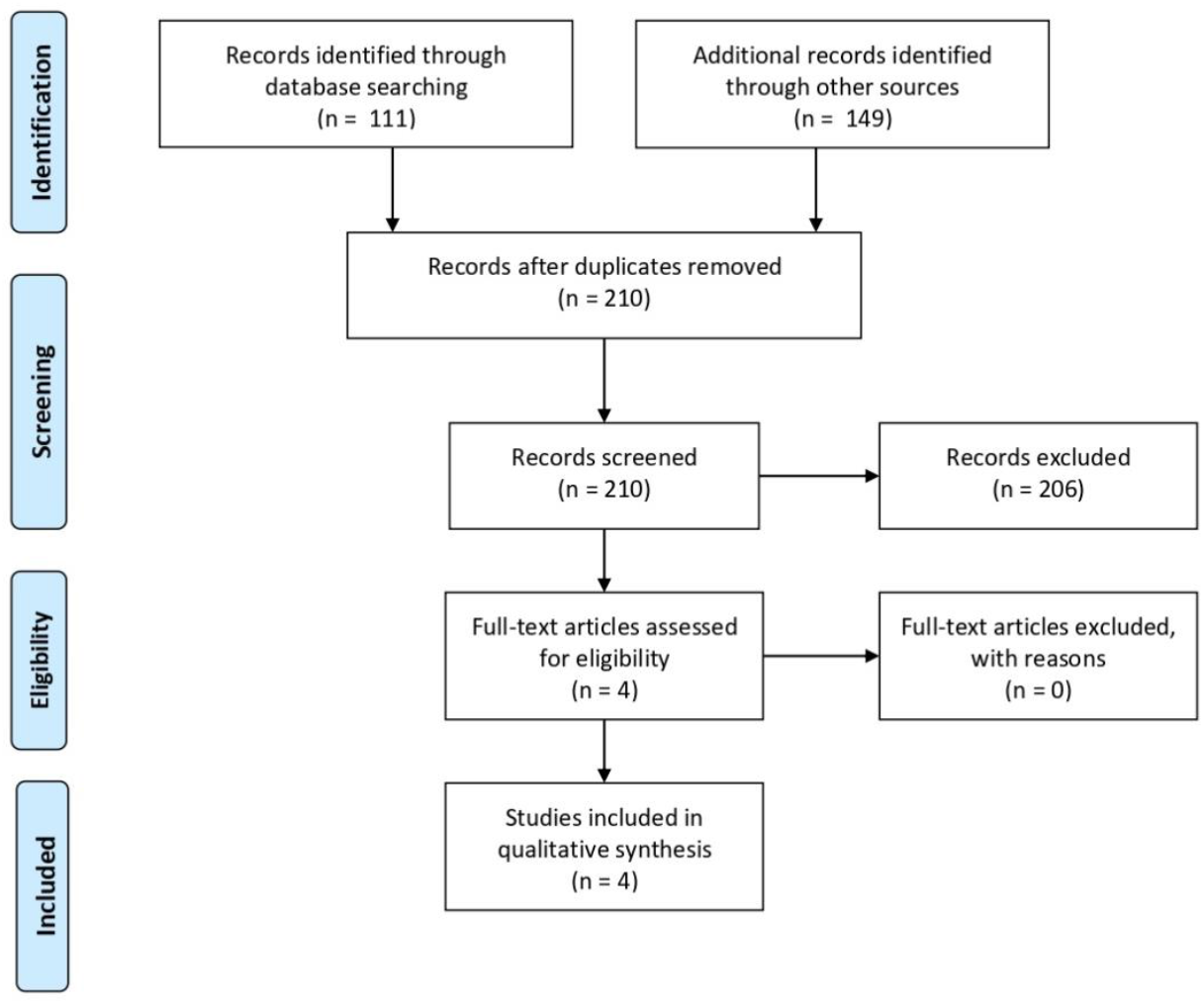
PRISMA Flow Diagram for included studies in the current systematic review

Three out of the total of four studies were by Molsted et al, and examined the effects of a 16-week supervised progressive high intensity dynamic resistance training (21–23). The focus of their two studies (2014), was mainly the relationship between hormones and other factors evaluated using blood samples with muscle performance and fiber size from muscle biopsies. However, differences in fiber size between baseline and after training were also reported and included in the current review. The focus of Molsted et al 2015 (24) was muscle specific with results related to fiber size, fiber type, satellite cells and myonuclei before and after the training period. It should be mentioned that the data of the above three studies were obtained as part of their previous study (25). The fourth study by Lewis et al 2015 (26) examined the effects of different types of exercise (endurance, strength or combination) for a 18-week period on muscle characteristics. All muscle biopsies were from their previous study assessing metabolic and morphometric profile of HD patients (27). Moreover, Lewis et al (26) compared the results not only between baseline and after training period but also between healthy control individuals and HD patients. Specifically, control subjects did not participate in any exercise training protocol while HD patients were separated in 4 groups: one group of HD patients did not participate in any exercise training protocol. The rest of the patients were enrolled in endurance exercise training or strength training or combination of endurance and strength training. All the four studies examined muscle biopsies from vastus lateralis muscle of dialysis patients.

The results were divided in 5 categories according to the focus of the muscle biopsy analysis: Cross Sectional Area (4 studies), fiber type (2 studies), satellite cells/myonuclei (1 study), mitochondrial markers (1 study) and capillaries (1 study).

## ANALYSIS AND DISCUSSION

### Cross Sectional Area (CSA)

Muscle fiber area is an important determinant of force generation (28) and it has been evaluated in many studies investigating uremic myopathy. It has been well established that CKD is accompanied by muscle atrophy with observations of smaller fiber cross-sectional areas (CSA) compared to healthy individuals. The latter has been reported in all muscle fiber types but mainly in fast-twitch fibers (type II) (8–10). All the 4 studies of the current review examined the effect of exercise training on fiber CSAs with controversial results. Specifically, Molsted et al, in three studies (21–23), did not report any significant difference in CSA values after the 16-week high intensity dynamic resistance training. On the other hand, Lewis et al (26), reported that type I and type IIA single muscle fibers’ CSAs were significantly reduced by 10% in patients enrolled at the exercise training program which combined strength and endurance training **(Table 1)**.

**Table 1.** Effects of exercise training on fiber cross sectional areas

| Title | Authors | Participants | Muscle | Intervention | Cross Sectional Area (CSA) |
| --- | --- | --- | --- | --- | --- |
| Interleukin-6 and vitamin D status during high-intensity resistance training in patients with chronic kidney disease | Molsted, Eiken et al 2014 | 36 dialysis patients (30HD/6PD) [age: 54 ± 2 (SEM)] | vastus lateralis | 16-week supervised progressive high-intensity dynamic resistance training 3 times weekly (N=23) | **Type I fibers:** pre-training: 4216 ± 311; post-training: 4427 ± 392; **Type II fibers:** pre-training: 3185 ± 233; post-training: 3606 ± 275 (values are in $\mu\text{m}^2$ MEAN±SEM) |
| Resistance Training and Testosterone Levels in Male Patients with Chronic Kidney Disease Undergoing Dialysis | Molsted, Andersen et al 2014 | 20 male dialysis patients (19HD/1PD) [age: 65 (26-74)] | vastus lateralis | 16-week supervised progressive high-intensity dynamic resistance training 3 times weekly (N=12) | **Type I fibers:** pre-training: 4760 (2853-7891); post-training: 4730 (2273-9204); **Type II fibers:** pre-training: 3246 (2284-5336); post-training: 3832 (2829-6953) [values are in $\mu\text{m}^2$ Median (range)] |
| Fiber type-specific response of skeletal muscle satellite cells to high-intensity resistance training in dialysis patients | Molsted et al 2015 | 21 dialysis patients (18HD/3PD) [age: 54 ± 3 (SEM)] | vastus lateralis | 16-week supervised progressive high-intensity dynamic resistance training 3 times weekly (N=21) | **Type I fibers:** pre-training: 4502±304; post-training: 4734±386; **Type II fibers:** pre-training: 3189±242; post-training: 3635±317 (values are in $\mu\text{m}^2$ MEAN±SEM) |
| Effect of endurance and/or strength training on muscle fiber size, oxidative capacity, and capillarity in hemodialysis patients | Lewis et al 2015 | a) 22 normal controls (age: $40.9 \pm 2.6$) b) 59 HD patients (age: $42.1 \pm 1.5$): b1) 15 HD patients/no training (NT); b2) 13 HD patients/endurance training (ET); b3) 17 HD patients/strength training (ST); b4) 14 HD patients/endurance & strength training (EST) (SEM) | vastus lateralis | 18-week supervised b2)endurance training (N=13), b3) strength training (N=17) b4) combination of endurance and strength training (N=14) [ST immediately before HD, ET during HD]; [a (N=22) and b1 (N=15) no training] | **Type I fibers:** a) pre-training: ~3900; post-training: ~3900; b1) pre-training: ~5000; post-training: ~5200; b2) pre-training: ~4800; post-training: ~5000; b3) pre-training: ~5500; post-training: ~5200; b4) pre-training: $5223 \pm 440$; post-training: $\downarrow 4683 \pm 370$; **Type IIA fibers:** a) pre-training: ~3750; post-training: ~3700; b1) pre-training: ~4700; post-training: ~4700; b2) pre-training: ~4500; post-training: ~4800; b3) pre-training: ~5000; post-training: ~4700; b4) pre-training: $4749 \pm 422$; post-training: $\downarrow 4291 \pm 387$; **Type IIX fibers:** a) pre-training: ~3300; post-training: ~3200; b1) pre-training: ~4250; post-training: ~4250; b2) pre-training: ~4100; post-training: ~4250; b3) pre-training: ~4500; post-training: ~4250; b4) pre-training: ~4100; post-training: ~3800 (values are in $\mu\text{m}^2$ MEAN±SEM) |

↑↓: significant difference compared to baseline values, ~: values were estimated according to figures; ± standard error of the mean (SEM), HD stands for hemodialysis, PD stands for peritoneal dialysis.

Therefore, three studies (21–23) reported no differences after a training period with resistance exercise and one study (26) showed significantly smaller CSAs of type I and IIA fibers CSAs after a period of strength and endurance training. However, older studies have reported different results. For example, Sakkas et al (29) who studied the effect of a six-month aerobic exercise on gastrocnemius muscle biopsies of dialysis patients reported significant increases of CSA values after the training period by 32%, 54% and 36% for type I, IIA and IIX fibers respectively. Similarly, Castaneda et al (30) who examined the effect of a 12-week resistance exercise training and low protein diet on vastus lateralis muscle biopsies reported significant increase of type I and II CSAs compared to low protein diet alone. Moreover, it should be mentioned that Lewis et al (26) who reported reduction of fiber areas after the training period, had previously reported larger CSA baseline values in HD patients compared to controls (26). However, most of the studies so far have reported muscle atrophy in CKD, both in pre-dialysis and dialysis patients. For example, Sakkas et al (3) who examined fibers’ CSAs in rectus abdominis muscle biopsies from pre-dialysis patients reported smaller CSAs in patients than in controls by 15, 26 and 28% for type I, IIA and IIA, respectively. Moreover, Johansen et al (31) who studied the lower leg CSA using MRI did not report significant differences in total muscle CSA but significant atrophy in contractile CSA of HD patients compared to controls.

### Fiber type

A muscle fiber consists of hundreds or thousands of myofibrils which are the contractile structures of a muscle cell (32). Each myofibril consists of many sarcomeres, each one consisting of a thick and a thin filament; the thick filaments consist mainly of myosin and the thin filaments mainly consist of actin, troponin and tropomyosin molecules. According to the myosin heavy chain (MHC) isoforms’ expression, fast fibers have been categorized into three main subcategories known as IIA, IIX and IIB (IIB are not expressed in humans). Except type I which is the slowest fiber type, type IIA is the slowest form of fast fibers and it is followed by IIX and IIB which are the fastest types (33,34). Many studies so far have reported modifications in the expression of MHC and therefore different muscle fiber proportions in muscle biopsies of CKD patients compared to healthy individuals. In the current systematic review two studies examined the effect of exercise training on fiber type distribution with similar results. Specifically, Molsted at al (24) reported no differences in distribution of type I nor of type II muscle fibers after the exercise training period. Similarly, Lewis et al 2015 who examined more MHC isoforms (I, IIA and IIX) reported no differences after the training period **(Table 2)**.

**Table 2.** Effects of exercise training on fiber type

| Title | Authors | Participants | Muscle | Intervention | Fiber type |
| --- | --- | --- | --- | --- | --- |
| Fiber type-specific response of skeletal muscle satellite cells to high-intensity resistance training in dialysis patients | Molsted et al 2015 | 21 dialysis patients (18HD/3PD) [age: 54 ± 3 (SEM)] | vastus lateralis | 16-week supervised progressive high-intensity dynamic resistance training 3 times weekly (N=21) | **Type I fibers:** pre-training: 45.4±3.2; post-training: 47.1±3.4; **Type II fibers:** pre-training: 54.6±3.2; post-training: 52.9±3.4 (values are in % MEAN±SEM) |
| Effect of endurance and/or strength training on muscle fiber size, oxidative capacity, and capillarity in hemodialysis patients | Lewis et al 2015 | a) 22 normal controls (age: 40.9 ± 2.6) b) 59 HD patients (age: 42.1 ± 1.5): b1) 15 HD patients/no training (NT); b2) 13 HD patients/endurance training (ET); b3) 17 HD patients/strength training (ST); b4) 14 HD patients/endurance & strength training (EST) (SEM) | vastus lateralis | 18-week supervised b2)endurance training (N=13), b3) strength training (N=17) b4) combination of endurance and strength training (N=14) [ST immediately before HD, ET during HD]; [a (N=22) and b1 (N=15) no training] | **Type I fibers:** a) pre-training: ~39; post-training: ~37; b1) pre-training: ~41; post-training: ~40.5; b2) pre-training: ~40; post-training: ~39.5; b3) pre-training: ~39; post-training: ~38.5; b4) pre-training: ~40; post-training: ~38.5; **Type IIA fibers:** a) pre-training: ~42; post-training: ~41; b1) pre-training: ~41; post-training: ~40; b2) pre-training: ~41.5; post-training: ~41; b3) pre-training: ~42; post-training: ~40.5; b4) pre-training: ~42; post-training: ~41.5; **Type IIX fibers:** a) pre-training: ~19.5; post-training: ~22; b1) pre-training: ~17.5; post-training: ~19; b2) pre-training: ~17.5; post-training: ~19; b3) pre-training: ~18; post-training: ~20; b4) pre-training: ~17; post-training: ~18 (values are in % MEAN±SEM) |

↑↓: significant difference compared to baseline values, ~: values were estimated according to figures; ± stands for standard error of the mean (SEM), HD stands for hemodialysis, PD stands for peritoneal dialysis.

Therefore, both studies agree that the training period did not result in alterations of fiber type distribution. These results are in accordance with an older study by Sakkas et al (29) who studied the effect of a six-month aerobic exercise on gastrocnemius muscle biopsies of dialysis patients and also reported no changes in MHC distribution after the training period. Apart from the above findings, it has to be mentioned that Lewis et al (26) who reported no alterations in fiber type distribution after the training period, had previously reported that HD patients had a significantly higher proportion of type I fibers and lower proportion of type IIX compared to controls (27). According to the authors the higher proportion of type I fibers may act as an adaptive response in order to maintain the oxidative capacity. However, older studies have reported different results. For example, Molsted et al (11) found significantly lower relative distribution of MHCI (type I) in HD patients (∼35% vs ∼59%) compared to controls in vastus lateralis muscle and significantly higher MHCIIX (type IIX) in HD patients (∼29% vs ∼7%) compared to controls. As it is discussed by the authors, an increase in fibers expressing MHCIIX has been linked to physical inactivity in healthy individuals and the physical inactivity which has been observed in HD patients probably leads to the higher content of this MHC isoform. The histochemical assessments of Fahal et al (10) also revealed a lower prevalence of type I (by 10%) and higher prevalence of type IIA (by 4%) and IIB (by 6%) fibers in muscle biopsies from the right quadriceps of dialysis patients when compared to healthy controls but without any significant difference between groups. On the other hand, in the study of Crowe et al (8) who examined muscle fibers’ type using staining for ATPase activity there was not found any significant difference between HD and control subjects in the proportion of type I nor of type II fibers excised from quadriceps femoris muscles. Similarly, Wagner et al (9) who examined rectus femoris muscle biopsies from young HD patients compared to healthy controls did not observe any difference in fiber type distribution between groups.

### Satellite cells/ myonuclei

Satellite cells (SC) and myonuclei are integrally involved in proliferative and regenerative response of skeletal muscle to any stimuli such as atrophy and exercise (35,36). SCs are located between the sarcolemma and basal lamina of muscle fibers and they are the main source of myonuclear addition (36). During aging and disease, the number of SCs is reduced while resistance exercise training is associated with increased number of SCs (37). Under normal conditions, resistance exercise training activates SC proliferation and promotes muscle hypertrophy (37). The effects of resistance exercise training on satellite cells of CKD patients’ muscle biopsies was studied only by Molsted et al (24). Apart from the SC content, this study also examined the myonuclear number and domain (the area of cytoplasm which is controlled by a single myonucleus). Muscle atrophy and disease have been associated with loss of myonuclei and reduction of myonuclear domain size, while hypertrophy has been associated with increased number of myonuclei without changes in myonuclear domain size (38). Molsted at al 2015 (21), examined the effect of resistance training on satellite cells’ (SC) content, myonuclear number and myonuclei domain. After the training period, SC content was increased in type I fibers without any change in myonuclear number and domain. On the other hand, in type II fibers there was not observed any significant difference in SC content and myonuclear domain after the training period but a significant increase in myonuclear number by ∼12% **(Table 3)**.

**Table 3.** Effects of exercise training on satellite cells/myonuclei

| Title | Authors | Participants | Muscle | Intervention | SC content | Myonuclear number | Myonuclear domain |
| --- | --- | --- | --- | --- | --- | --- | --- |
| Fiber type-specific response of skeletal muscle satellite cells to high-intensity resistance training in dialysis patients | Molsted et al 2015 | 21 dialysis patients (18HD/3PD) [age: $54 \pm 3$ (SEM)] | vastus lateralis | 16-week supervised progressive high-intensity dynamic resistance training 3 times weekly (N=21) | **Type I fibers:** a) pre-training: $0.053 \pm 0.006$ b) post-training: $\uparrow 0.061 \pm 0.005$; **Type II fibers:** a) pre-training: $0.044 \pm 0.00$ b) post-training: $0.035 \pm 0.003$ (values are SCs/fiber MEAN±SEM) | **Type I fibers:** a) pre-training: $1.90 \pm 0.12$ b) post-training: $2.07 \pm 0.12$; **Type II fibers:** a) pre-training: $1.85 \pm 0.10$ b) post-training: $\uparrow 2.09 \pm 0.11$ (values are number/fiber MEAN±SEM)(N=16) | **Type I fibers:** a) pre-training: $2411 \pm 174$ b) post-training: $2353 \pm 124$; **Type II fibers:** a) pre-training: $1785 \pm 174$ b) post-training: $1683 \pm 138$ (values are in $\mu\text{m}^2$ MEAN±SEM)(N=16) |

$\uparrow\downarrow$ : significant difference compared to baseline values, ± stands for standard error of the mean (SEM), HD stands for hemodialysis, PD stands for peritoneal dialysis.

To our knowledge this study was the first one to study the effects of resistance training on satellite cells and myonuclei of CKD muscle biopsies and the results should be confirmed and further investigated in future studies. Resistance exercise training in healthy individuals has been associated with changes in both SC and myonuclear content and with parallel increase in CSA of muscle fibers (39). However, in the study of Molsted et al 2015 (21) there were not reported in parallel alterations of SC and myonuclear content nor any alterations in fibers CSAs. It should be also mentioned that a recent study by Brightwell et al (19) found that the distance between capillaries and satellite cells in vastus lateralis muscle biopsies of CKD patients (dialysis and non-dialysis) was significantly greater compared to control muscle biopsies indicating impaired SC activity and function in CKD patients.

### Mitochondrial markers

Muscle contraction requires energy and mitochondria are the key organelles for energy production in the form of ATP. Mitochondrial form, content and function may be impaired due to many clinical conditions (40). Indeed, recent studies have reported significant mitochondrial abnormalities in muscle biopsies of both dialysis and pre-dialysis CKD patients (12,13). However, only one study was found to examine the effect of exercise training on mitochondrial markers of CKD human muscle biopsies. Specifically, Lewis et al 2015 (26) investigated the effect of exercise training on succinate dehydrogenase (SDH) activity, which is a key mitochondrial enzyme with crucial role in energy metabolism and an indicator of mitochondrial function. That study reported that SDH activity was not significantly affected by any type of exercise training. However, SDH activity was found to be increased at the endurance exercise group in IIA and IIX fibers by 16.3% and 19.6% respectively. Similarly SDH activity was found to be increased at the strength training group in type IIA fibers by 8.9% **(Table 4)**.

**Table 4.** Effects of exercise training on mitochondrial markers

| Title | Authors | Participants | Muscle | Intervention | SDH activity |
| --- | --- | --- | --- | --- | --- |
| Effect of endurance and/or strength training on muscle fiber size, oxidative capacity, and capillarity in hemodialysis patients | Lewis et al 2015 | a) 22 normal controls (age: $40.9 \pm 2.6$) b) 59 HD patients (age: $42.1 \pm 1.5$): b1) 15 HD patients/no training (NT); b2) 13 HD patients/endurance training (ET); b3) 17 HD patients/strength training (ST); b4) 14 HD patients/endurance & strength training (EST) (SEM) | vastus lateralis | 18-week supervised b2)endurance training (N=13), b3) strength training (N=17) b4) combination of endurance and strength training (N=14) [ST immediately before HD, ET during HD]; [a (N=22) and b1 (N=15) no training] | **Type I fibers:** a) pre-training: ~2.4; post-training: ~2.6; b1) pre-training: ~1.65; post-training: ~1.6; b2) pre-training: ~1.6; post-training: ~1.65; b3) pre-training: ~1.75; post-training: ~1.6; b4) pre-training: ~1.75; post-training: ~1.75; **Type IIA fibers:** a) pre-training: ~2; post-training: ~1.8; b1) pre-training: ~1.15; post-training: ~1.25; b2) pre-training: ~1.15; post-training: ~1.3; b3) pre-training: ~1.15; post-training: ~1.25; b4) pre-training: ~1.15; post-training: ~1.25; **Type IIX fibers:** a) pre-training: ~1.9; post-training: ~1.8; b1) pre-training: ~1.0; post-training: ~1.0; b2) pre-training: ~1.0; post-training: ~1.25; b3) pre-training: ~1.1; post-training: ~1.1; b4) pre-training: ~1.1; post-training: ~1.1 (values are in mM Fumarate/L/min MEAN±SEM) |

↑↓: significant difference compared to baseline values, ~: values were estimated according to figures; ± stands for standard error of the mean (SEM), HD stands for hemodialysis, PD stands for peritoneal dialysis.

These results indicate that exercise training could potentially improve mitochondrial function in CKD skeletal muscle. However, in an older study, Sakkas et al (29) who investigated the effects of aerobic exercise training on gastrocnemius muscle biopsies of dialysis patients reported that another indicator of mitochondrial function, cytochrome c oxidase concentration, was not altered after the six-month training period. On the other hand, Balakrishnan et al (41), who investigated the effect of a twelve-week resistance exercise training on mitochondrial biogenesis of vastus lateralis muscle biopsies from pre-dialysis patients reported a significant increase in median mtDNA copy number by 7.5% compared to baseline. Additionally, the changes in mtDNA copy number were significantly associated (positively) with alterations in CSAs of both fiber types. Thus, resistance training improved mitochondrial content and muscle mass of patients.

### Capillaries

Capillaries play a crucial role in muscle function and any abnormality in capillary number, capillary to fiber ratio (i.e. number of capillaries/number of fiber in a muscle section) and capillary density (capillaries/mm^2^) can impair energy production and oxygen delivery in muscle tissue (42). Moreover, capillaries interact with satellite cells (SC) for their proper activation and function as well as for angiogenesis. Alterations in the distance between satellite cells and capillaries can also affect this interaction (43). Although capillaries can be negatively affected during aging and disease (44), only one recent study was found to investigate the effect of exercise training on capillarity of HD patients’ muscle biopsies. Specifically, Lewis et al (26), reported that endurance exercise increased the number of capillaries per individual fiber in type I and type IIA fibers by 6% and 4% respectively. Furthermore, combination of endurance and strength training increased the number of capillaries per individual fiber in type I, type IIA and type IIX fibers by 7%, 4% and 3% respectively. Independently of fiber type, it was also found that endurance training and the combination of endurance and strength training led to a significant increase of capillary to fiber ratio by 9.6% and 11% respectively. The capillary density was also significantly increased after endurance training by 14.3% and after combination of endurance and strength training by 28% **(Table 5)**.

**Table 5.** Effects of exercise training on capillaries

| Authors | Participants | Muscle | Intervention | Capillaries |
| --- | --- | --- | --- | --- |
| Lewis et al 2015 | a) 22 normal controls (age: 40.9 ± 2.6) b) 59 HD patients (age: 42.1 ± 1.5): b1) 15 HD patients/no training (NT); b2) 13 HD patients/endurance training (ET); b3) 17 HD patients/strength training (ST); b4) 14 HD patients/endurance & strength training (EST) (SEM) | vastus lateralis | 18-week supervised b2)endurance training (N=13), b3) strength training (N=17) b4) combination of endurance and strength training (N=14) [ST immediately before HD, ET during HD]; [a (N=22) and b1 (N=15) no training] | **Capillaries per fiber: Type I fibers:** a) pre-training: ~4.3; post-training: ~4.3; b1) pre-training: ~4.6; post-training: ~4.8; b2) pre-training: ~4.6; post-training: ↑~4.9; b3) pre-training: ~4.6; post-training: ~4.7; b4) pre-training: ~4.5; post-training: ↑~4.9; **Type IIA fibers:** a) pre-training: ~4.3; post-training: ~4.3; b1) pre-training: ~4.7; post-training: ~4.9; b2) pre-training: ~4.7; post-training: ↑~4.9; b3) pre-training: ~4.7; post-training: ~4.7; b4) pre-training: ~4.7; post-training: ↑~4.9; **Type IIX fibers:** a) pre-training: ~3.9; post-training: ~3.9; b1) pre-training: ~4.8; post-training: ~4.9; b2) pre-training: ~4.8; post-training: ~4.8; b3) pre-training: ~4.8; post-training: ~4.8; b4) pre-training: ~4.7; post-training: ↑~4.8 (values are capillaries per individual fiber MEAN±SEM) **Capillary to fiber ratio** a) pre-training: ~1.65; post-training: ~1.7; b1) pre-training: ~1.8; post-training: ~1.85; b2) pre-training: ~1.75; post-training: ↑~1.95; b3) pre-training: ~1.8; post-training: ~1.9; b4) pre-training: ~1.75; post-training: ↑~2 (values are number of capillaries per individual fiber MEAN±SEM) **Capillary density** a) pre-training: ~540; post-training: ~560; b1) pre-training: ~370; post-training: ~350; b2) pre-training: ~340; post-training: ↑~390; b3) pre-training: ~370; post-training: ~330; b4) pre-training: ~310; post-training: ↑~400 (values are capillaries per mm<sup>2</sup> MEAN±SEM) |

↑↓: significant difference compared to baseline values, ~: values were estimated according to figures; ± stands for standard error of the mean (SEM), HD stands for hemodialysis, PD stands for peritoneal dialysis.

The above results indicate that exercise training may improve muscle capillarity in CKD. Similarly, in an older study, Sakkas et al (29) who investigated the effect of a six-month aerobic exercise training in skeletal muscle morphology of dialysis patients found that capillary contact per fiber was also improved by 24% in gastrocnemius muscle biopsies. However, it has to be mentioned that in their companion study, Lewis et al (27) reported that the number of capillaries contacting each fiber was significantly higher in HD patients compared to controls by 9%, 10% and 23% for type I, IIA and IIX fibers respectively. Additionally, capillary/fiber ratio was significantly higher in HD patients by 11% but capillary density was significantly lower by 34% in HD patients. However, as previously mentioned, in contrast to other studies, this study also reported larger fiber CSAs in muscle fibers of CKD muscle.

Therefore, according to the authors, although capillaries per fiber and capillary/fiber ratio was found to be increased in HD patients, their larger fibers’ CSAs resulted to low capillary density and therefore insufficient oxygen supply especially under conditions with high oxygen demands such as exercise.

### Effects of exercise training on muscle performance

Apart from the results from the analysis of muscle biopsies, it has to be reported that in all the three studies of Molsted, muscle function parameters of patients were also tested before and after the training period with reports of significant improvements in muscle performance. Specifically in the study of Molsted et al 2014 (23), leg extension power was significantly increased after the training period by 23-25%. Physical function measured through the chair stand test was also significantly improved after training period by ∼21%. Molsted et al 2014 (22) also reported that muscle strength measured by knee extension was significantly improved after training period by 19-25%. Lastly, Molsted et al 2015 (21) reported that after the training period, muscle strength (torque associated with knee extension) was significantly increased by 20-25%. Moreover, in the study of Lewis et al (26), although the data about the effect of exercise training are not reported, in their companion study (27), they observed significant improvements in strength and endurance of HD patients **(Table 6)**.

**Table 6.** Effects of exercise training on muscle performance

| Title | Authors | Participants | Muscle | Intervention | Muscle performance |
| --- | --- | --- | --- | --- | --- |
| Interleukin-6 and vitamin D status during high-intensity resistance training in patients with chronic kidney disease | Molsted, Eiken et al 2014 | 36 dialysis patients (30HD/6PD) [age: 54 ± 2 (SEM)] | vastus lateralis | 16-week supervised progressive high-intensity dynamic resistance training 3 times weekly (N=23) | left leg a) pre-training: 2.01 ± 0.21 b)post-training: ↑2.48 ± 0.20; right leg a) pre-training: 2.11 ± 0.20 b)post-training: ↑2.64 ± 0.19 (power: values in W/kg MEAN±SEM) |
| Resistance Training and Testosterone Levels in Male Patients with Chronic Kidney Disease Undergoing Dialysis | Molsted, Andersen et al 2014 | 20 male dialysis patients (19HD/1PD) [age: 65 (26-74)] | vastus lateralis | 16-week supervised progressive high-intensity dynamic resistance training 3 times weekly (N=12) | left leg a) pre-training:341 (193-558) b)post-training: ↑400 (238-657); right leg a) pre-training: 349 (200-583) b)post-training: ↑460 (258-695)[strength values in N median (range)] |
| Fiber type-specific response of skeletal muscle satellite cells to high-intensity resistance training in dialysis patients | Molsted et al 2015 | 21 dialysis patients (18HD/3PD) [age: 54 ± 3 (SEM)] | vastus lateralis | 16-week supervised progressive high-intensity dynamic resistance training 3 times weekly (N=21) | left leg a) pre-training: 102 ± 9 b)post-training: ↑123 ± 11; right leg a) pre-training: 111 ± 10 b)post-training: ↑139 ±13 (torque values in Nm MEAN±SEM) |

↑↓: significant difference compared to baseline values, ± stands for standard error of the mean (SEM), HD stands for hemodialysis, PD stands for peritoneal dialysis.

## CONCLUSIONS

To our knowledge, this is the first systematic review on the effects of counteracting measures on muscle biopsies of CKD patients of the last decade. Among other non-pharmaceutical and pharmaceutical counteracting measures, only exercise interventions were found to be examined in the last ten years. The effect of exercise training on muscle biopsies of CKD patients was studied only in 4 studies and the reported muscle contraction determinants were the following: Cross sectional area, fiber type, satellite cells/myonuclei, mitochondrial markers and capillaries.

From the current systematic review it can be concluded that exercise intervention, clearly improves muscle performance of patients but the results from muscle biopsies are limited and frequently controversial. Specifically, although the presented studies agree that exercise training does not result in fiber type alterations and that muscle capillarity is improved after exercise training, the effects of exercise training on fiber cross sectional area and mitochondrial markers remain controversial. Moreover, to our knowledge, the effects of exercise training on satellite cells and myonuclei has been studied only in one study so far and the results should be further confirmed and investigated. The controversial results could be due to the fact that CKD is associated with a host of comorbidities such as diabetes mellitus and corresponding pharmaceutical treatments which could affect the results of muscle contraction determinants. Other factors such as exercise type, stage of the disease and methodological approach could be also implicated.

It can be also concluded that although uremic myopathy is a devastating condition in CKD patients, in recent years only a few studies have examined the effects of counteracting measures on muscle biopsies of CKD patients, and they are limited to exercise interventions. However, a host of counteracting measures have shown to improve muscle performance in CKD patients or animal models and could be also assessed at the muscle biopsy level of CKD patients (alone or in combination with other interventions). For example, a recent study (45) with iron supplementation in an animal model of CKD showed significant improvements in muscle contraction properties of single muscle fibers indicating that timely maintenance of iron balance in CKD patients could also maintain or improve skeletal muscle quality and quantity. Therefore, future studies could verify the results in muscle biopsies of human patients (preferably during catheter insertion or transplantation to avoid further discomfort of patients). An additional parameter which should be considered is that most of the studies so far have been conducted in dialysis patients. However, interventional studies in pre-dialysis patients could delay/ameliorate the negative effects of the disease on skeletal muscle and reveal further insights into the progression of uremic myopathy. Moreover, it should be stated that the focus of the interventional studies of this review were not always skeletal muscle per se and the assessed muscle contraction determinants were mainly limited to cross sectional areas, fiber type alterarations, capillaries and mitochondrial markers. However, there is evidence that skeletal muscle of CKD patients presents with a number of additional abnormalities such as oxidative/nitrosative stress (14,15), inflammation (16,17), fat accumulation/infiltration (16,17) and fibrosis (18,19) which can also affect muscle contraction. Therefore, in order to better understand the effects of interventional studies on skeletal muscle of CKD patients, future studies should examine their effects on those parameters as well. The above will help us to support the design of the most promising interventions for skeletal muscle quality and quantity with a potential of better understanding the mechanisms underlying uremic myopathy aiming to delay or alleviate its devastating impacts.

## Data Availability

All data produced in the present work are contained in the manuscript.

## ACKNOWLEDGMENTS

- GM thanks Miss Virginia Smixioti (MSc) for the literature search.
- GM was supported in the context of the project ‘Reinforcement of Postdoctoral Researchers - 2nd Cycle’ (MIS-5033021), co-financed by Greece and the European Union (European Social Fund - ESF) through the Operational Programme «Human Resources Development, Education and Lifelong Learning», implemented by the State Scholarships Foundation (IKY).

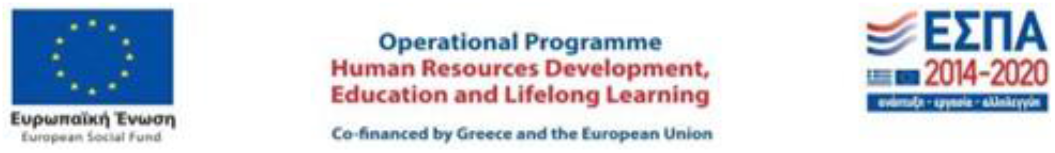

